# The Role of Early Cerebral Edema and Hematoma assessment in Aneurysmal Subarachnoid Hemorrhage (a-SAH) in predicting Structural Brain Abnormalities in Cognitive Impairments

**DOI:** 10.1101/2023.12.05.23299565

**Authors:** Ming-Dong Wang, Qian-Hui Fu, Andrew Ni, Yun-Peng Yuan, Chun-Hui Li, Zhan-Xiang Wang, Hong Wang

## Abstract

**Background:** Early assessment and management of cerebral edema and hematoma following aneurysmal subarachnoid hemorrhage (a-SAH) can significantly impact clinical cognitive outcomes. However, current clinical practices lack predictive models to identify early structural brain abnormalities affecting cognition. To address this gap, we propose the development of a predictive model termed the a-SAH Early Brain Edema/Hematoma Compression Neural (Structural Brain) Networks Score System (SEBE-HCNNSS).

**Methods:** In this study, 202 consecutive patients with spontaneous a-SAH underwent initial computed tomography (CT) or magnetic resonance imaging (MRI) scans within 24 hours of ictus with follow-up 2 months after discharge. Using logistic regression analysis (univariate and multivariate), we evaluated clinically relevant factors and various traditional scale ratings for cognitive impairment (CI). Risk factors with the highest area under the curve (AUC) values were included in the multivariate analysis and least absolute shrinkage and selection operator (LASSO) analysis or Cox regression analysis.

**Results:** A total of 177 patients were enrolled in the study, and 43 patients were classified with a high SEBE-HCNNSS grade (3 to 5). After a mean follow-up of 2 months, 121 individuals (68.36%) with a-SAH and 3 control subjects developed incident CI. The CT inter-observer reliability of the SEBE-HCNNSS scale was high, with a Kappa value of 1. Furthermore, ROC analysis identified the SEBE-HCNNSS scale (OR 3.322, 95% CI 2.312-7.237, p = 0.00025) as an independent predictor of edema, CI, and unfavorable prognosis. These results were also replicated in a validation cohort.

**Conclusion:** Overall, the SEBE-HCNNSS scale represents a simple assessment tool with promising predictive value for CI and clinical outcomes post-a-SAH. Our findings indicate its practical utility as a prognostic instrument for risk evaluation after a-SAH, potentially facilitating early intervention and treatment.

## INTRODUCTION

The latest delineation of subgroups within the subarachnoid space provides a theoretical basis for more accurate diagnosis and treatment of subarachnoid hemorrhage (SAH)^1^. Specifically, aneurysmal subarachnoid hemorrhage (a-SAH) is a subtype of hemorrhage stroke associated with high morbidity and mortality, constituting a significant cause of neurological disease^1,2^. Common complications following a-SAH include early brain injury (EBI), symptomatic vasospasm, brain edema, delayed cerebral ischemia (DCI), delayed deficits, and cognitive impairments (CI)^3,4,5^. Despite advancements enhancing outcomes post-SAH, CI still affects 25% to 40% of stroke survivors^6,7^, and the precise risk factors and mechanisms underlying CI in SAH patients remain elusive.

Cognitive dysfunction can show substantial short-term variability within individuals^8^. Furthermore, in pathological conditions like a-SAH, structural brain abnormalities, such as cerebral edema and DCI, are strongly liked with the risk of developing CI^9,10^. Consequently, solely assessing cognitive performance can yield misleading conclusions^11^. At present, there is a lack of consensus on evaluation and detection tools for structural brain abnormalities (cerebral hematoma, edema) in CI. Although the Fisher scale on admission has been identified as an independent predictor for severe CI^12^, its correlation with structural brain abnormalities and subsequent CI, evaluated using Montreal Cognitive Assessment (MoCA) and Mini-Mental State Examination (MMSE), remains unknown.

Despite growing attention on EBI, defined as injury in the first 72 hours after SAH^13^, early evaluation of nerve fiber tracts has been overlooked. Typically, classification systems are grounded in accepted principles specific to their respective field. At present, a shortened version of MoCA has been used as the initial assessment tool for CI in patients through the recommendation of consensus conference guidelines^14^. However, this fails to capture the multifaceted nature of CI rooted in brain network functionalities. Cognitive processes, intricate and multifactorial, hinge on functional specificity, connectivity, brain architectonics, and topographic organization of brain areas^15,16,17^. Thus, functional brain network classification, delineating macro-scale networks, offers promise in understanding CI mechanisms. Six macro-scale functional brain networks have been delineated: the occipital network (ON), pericentral network (PN), dorsal frontoparietal network (D-FPN), lateral frontoparietal network (L-FPN), midcingulo-insular network (M-CIN), and medial frontoparietal network (M-FPN)^18^. Hence, this necessitates an alternative clinically useful marker for predicting CI using these macro-scale networks.

In this study, we aim to introduce a readily available, novel, radiographic scoring system, the a-SAH Early Brain Edema/Hematoma Compression Neural Networks Score System (SEBE-HCNNSS), to assess CI attributed to early brain damage and compression of neural networks. Modeled after the SEBES grade for early brain edema, the SEBE-HCNNSS is a semiquantitative CT grading scale. We hypothesize that this system not only accurately reflects the clinical grade of early brain edema post-SAH but also predicts the occurrence of CI and eventual clinical outcomes.

## MATERIALS AND METHODS

### Research Design and Study Population

The cognitive evaluation study enrolled 202 participants aged 18 to 70 diagnosed with a-SAH between Jan 2021 and Jan 2023. For this analysis, we used a case-cohort design. We excluded 25 participants who did not meet the predefined criteria, and the final analysis was performed on a cohort of 177 individuals. The study protocol was approved by the Institutional Review Board (Clinical Research Ethics Committee of the First Hospital of Hebei Medical University, ID: 2023-1001), and all participants and their proxies provided written informed consent.

Inclusion criteria: 1) first-ever stroke, diagnosed with SAH via CT scan within 24 –48 hours or lumbar puncture; 2) age between 18 and 70 years; 3) Confirmation of cerebral aneurysm by digital subtraction angiography (DSA) and/or CT angiography (CTA); 4) absence of neurological or psychiatric disease history and each unruptured intracranial aneurysm patient must be admitted to the hospital in excellent preoperative and pre-interventional condition; 5) Informed consent is signed by the patient and/or family.

Exclusion Criteria: 1) patients over 70 years old; 2) presence of neurological focal deficits or severe aphasia; 3) cognitive dysfunction or history of cognitive decline including craniotomy, antipsychotics, neurodegenerative diseases, and chronic subdural hematoma; 4) concurrent acute or chronic infections, corneal or pupillary abnormalities, severe autoimmune or systemic diseases such as rheumatic illnesses of the musculoskeletal system; 5) concurrent severe organic dysfunction; 6) patients with recurrent aneurysms or aneurysms not first diagnosed in our hospital; 7) diagnosis of major psychosis according to the Diagnostic and Statistical Manual of Mental Disorders, 4th Edition criteria^19^.

### Cognitive Assessment and Diagnosis Procedure

At the 2-month post-discharge mark, each participant underwent comprehensive neuropsychological testing using either MoCA or MMSE, which included a broad spectrum of cognitive domains such as construction, memory, attention, visual structural skills, language, abstract thinking, calculation, executive functions, psychomotor speed, and intellectual functioning. The MoCA scale ranges from 0 to 30 points, with 26–30 points classified as normal and <26 points classified as CI. The MMSE scale ranges from 0 to 30 points, with scores of 27–30 classified as normal, 21–26 as mild CI, 10–20 points as moderate CI, and 0–9 points as severe CI^20^. The MoCA scale was used to evaluate CI in the training cohort while the MMSE scale was used to evaluate CI in the validation cohort. The utilization of distinct cognitive assessment scales in the training and validation cohorts aimed to validate and enhance the robustness of findings across different cohorts and assessment tools, ensuring a comprehensive evaluation of cognitive impairment post-discharge.

### Clinical Variables

Clinical variables were selected based on previous recommendations which included general risk factors, baseline demographic data, and vascular risk factor data^1^. This included gender, age, blood pressure, lipid profile, glucose levels, smoking history, surgical time, aneurysm location and size, initial severity of bleed, clinical markers of EBI, SEBES scale^21^, Hunt-Hess scale^22^, modified Fisher grade, and Glasgow Coma Scale (GCS) at time of admission, and MoCA and MMSE at 2-months post-discharge (Details in online supplemental materials, Clinical Variables).

### Radiographic Variables and SEBES-HCNSS Criteria

The first available pretreatment and follow-up CT scan (within 24 hours of ictus) were reviewed by one of the authors (M.D.W) with fragment checking, manual correction, manual outlining, and insertion of identified cognitive networks following Fisher’s hierarchical classification and SEBES rules (Details in online supplemental materials, Radiographic Variables).

We defined SEBES-HCNNSS on a scale from 0 to 5 points. The SEBES-HCNSS score was determined by assessing the absence of visible sulci either because of effacement of sulci or loss of gray white differentiation at 2 predetermined levels in each hemisphere^21^: (1) Areas before the level of the insular cortex where the thalamus and basal ganglion are visible, including D-FPN, L-FPN, M-FPN and (2) areas of M-CIN in the periphery at the level of the lateral ventricle, including PN, ON and M-CIN (Table 1) (Details in online supplemental materials, Radiographic Variables).

**Table 1.** a-SAH Early Brain Edema/Hematoma Compression Neural Networks Score System (SEBE-HCNNSS) Criteria.

| Description of subarachnoid blood on cranial CT | Maximum Score |
| --- | --- |
| No visible blood, no effacement, and no widening of the longitudinal fissure of the brain. | 0 |
| Absence of visible sulci due to effacement, diffuse deposits, or thin layers of blood at specified brain locations (L-FPN or D-FPN or M-FPN or PN or ON or M-CIN or other), or widening of the longitudinal fissure. | 1 |
| Localized/diffuse blood deposits (<1 mm thick) at specified locations (L-FPN or D-FPN or M-FPN or PN or ON or M-CIN or other) in each section, without visible sulci in those areas at two predetermined levels in each hemisphere. | 2 |
| For specified locations (L-FPN or D-FPN or M-FPN or PN or ON or M-CIN or other) in each section, absence of visible sulci at two predetermined levels in each hemisphere or disruption of the grey-white matter junction, with blood pooling (<1 mm thick) in ventricles or cerebral pools (insular pools, circumferential pools, lateral fissure pools, interpeduncular pools, lateral ventricles). | 3 |
| Disappearance of sulci at two predetermined levels in each hemisphere or localized clots/thick blood layers ($\geq 1$ mm) within ventricles at specified brain locations (L-FPN or D-FPN or M-FPN or PN or ON or M-CIN or other). | 4 |
| Diffuse subarachnoid hemorrhage or absence of it with intracerebral/intraventricular blood clots at specified brain locations (L-FPN or D-FPN or M-FPN or PN or ON or M-CIN or other) | 5 |

### Localization of a-SAH

We also performed precise localization of the a-SAH (aneurysm location), as detected by CT, by manually tracing the hematoma or cerebral edema borders onto a template of brainstem nuclei in standard Montreal Neurological Institute space (The Harvard Ascending Arousal Network Atlas, www.martinos.org/resources/aan-atlas). Affected regions, including the L-FPN, PFCs, IFJ, IPL, M-CIN, and AI were delineated (Figure 1, online supplemental materials, Radiographic Variables).

**Figure 1.**
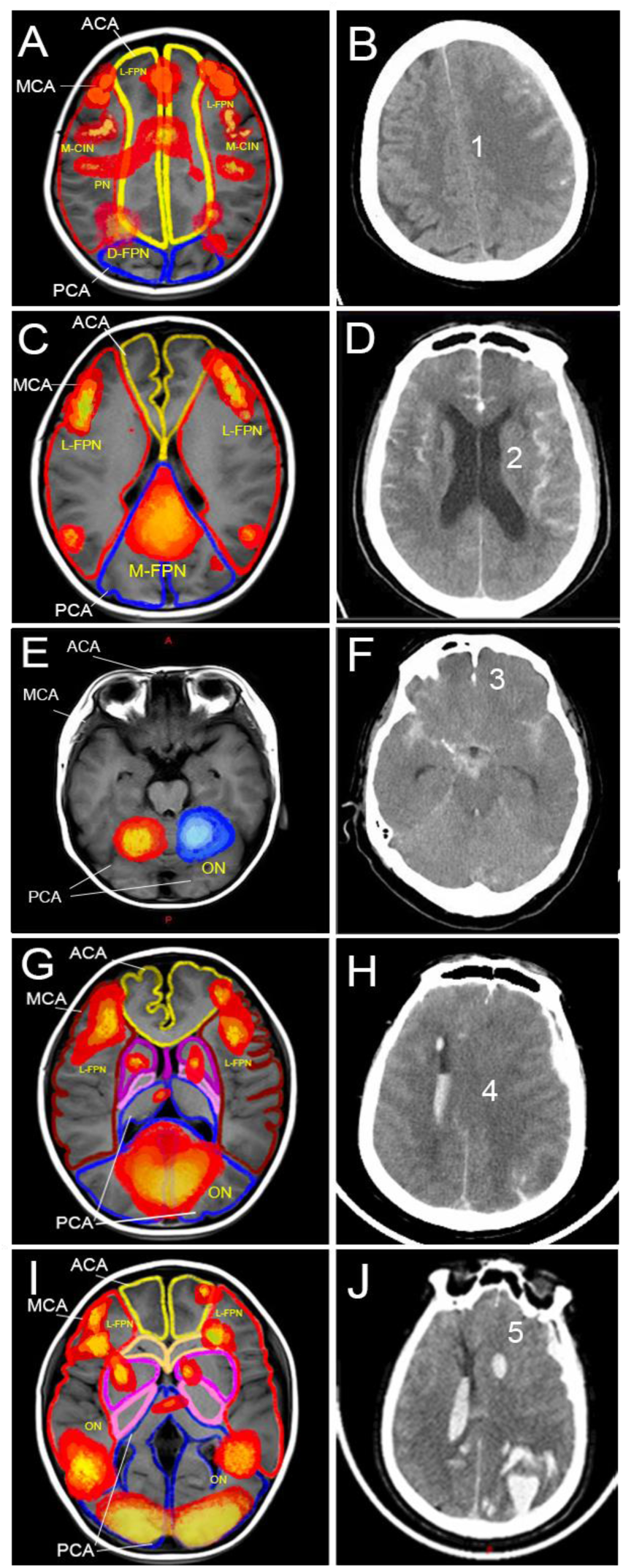
A, C, E G and I images show the distribution of different regions of cerebral arterial supply. Outlined in three different colors, yellow, red and blue. Yellow, red and blue refer to the broad cognitive domains with which a given anatomical system is most commonly associated. The B, D, F, H, J images show grade 5 of SEBE-HCNNSS with the effacement of sulci at 2 predetermined levels in each hemisphere, and the compression of cognitive network areas in the functional white matter regions.

### Validation cohort

A separate validation cohort comprised patients admitted with SAH subsequent to the initial cohort. The same inclusion criteria were applied to assess the prognostic value of SEBE-HCNNSS in predicting edema, hematoma, cognitive impairment, and unfavorable long-term prognosis.

### Statistical Analysis

Parametric screening of univariate analyses was conducted for the MoCA and MMSE score groups (training and validation sets) to identify significant differences and independent risk factors associated with CI. Multivariate regression models were used to explore correlations between independent risk factors and CI. Target variables were further identified using LASSO regression analyses. Six distinct brain structure regions were incorporated in the regression model to assess their ability to predict cognitive domain dependence and recurrence of vascular events. The accuracy of the SEBE-HCNNSS scale was evaluated against existing scales using receiver operating characteristic curves (ROCs), calibration plots, and dynamic component analysis/decision curve analysis (DCA) to ascertain its ability to predict the occurrence of edema, infarction and CI after SAH. This was done to determine which scale had the best combination of specificity and sensitivity so the best scale could be selected. We compared the models by determining the net categorical improvement rate and the combined discriminant improvement rate to select the best model. For the results of the MoCA and MMSE scale groups, we adjusted for location, size, Fisher grading, Hunt-Hess scale, EBES scale, GCS scale, and surgical approach. Cox proportional hazards models were used for all analysis phases. Statistical significance was determined by a two-tailed α of 0.05 in the first and second stages of analysis. In the third stage of analysis, Bonferroni adjustment was performed for multiple comparisons and an α level of 0.05 was used (Details in online supplemental materials, Statistical Analysis).

## RESULTS

### Participants characteristics

The study population consisted of 177 individuals meeting the inclusion criteria (67 male, 110 females, mean age 56.45±11.77 years), including aneurysm cases (a-SAH) and control subjects with unruptured intracranial aneurysms (UIA) (12 males, 13 females; mean age 53.04±15.60 years) (Table 2). Characteristics of the total study population and study population stratified by group are presented in Table 2. The mean age of the overall cohort was 57.01±10.98 years and 63.80% were female. Endovascular coiling was employed in 132 (74.58%) cases, while 37 (21.47%) cases underwent surgical clipping. The time from ictus onset to first CT was within 24 hours after admission. In addition, 148 (83.62%) ruptured aneurysms were in the anterior circulation and 29 (16.37%) ruptured aneurysms were in the posterior circulation. There were no significant differences between the two groups (p>0.05) across the different variables, except for time from onset to surgery, which was shorter in the a-SAH group (OR=238.286, 95%CI=29.933–1896.887, p<0.05). At the 2-month follow-up, 4 patients with UIA (16%) and 120 patients with a–SAH (78.43%) developed CI.

**Table 2.** Evaluation of Baseline Characteristics of 177 Participants in the Case-Cohort Study.

| Characteristics (Variable) | Participants (n=177) |  | Odds Ratio (95%CI) | p-value |
| --- | --- | --- | --- | --- |
|  | UIA (n=25) | a-SAH (n=152) |  |  |
| **Gender, male (37.85%)** |  |  | 1.645 (0.702–3.853) | 0.252 |
| Male | 12 (48%) | 55 (36.18%) |  |  |
| Female | 13 (52%) | 97 (63.825) |  |  |
| **Age, year** |  |  | 0.515 (0.183–1.452) | 0.21 |
| <65 | 20 (80%) | 53 (34.87%) |  |  |
| ≥65 | 5 (20%) | 99 (65.13%) |  |  |
| **Current Smoking (19.21%)** |  |  | 0.496 (0.14–1.759) | 0.278 |
| Smoking | 3 (12%) | 31 (20.39%) |  |  |
| Not Smoking | 22 (88%) | 121 (79.61%) |  |  |
| **SBP, mmHg** |  |  | 1.091 (0.46–2.584) | 0.843 |
| High | 14 (56%) | 88 (57.89%) |  |  |
| Normal | 11 (44%) | 64 (42.11%) |  |  |
| **LDL cholesterol, mmol/L** |  |  | 1.617 (0.587–4.452) | 0.353 |
| High | 6 (24%) | 25 (16.45%) |  |  |
| Normal | 19 (76%) | 127 (83.55%) |  |  |
| **Blood Sugar** |  |  | 1.344 (0.46–3.927) | 0.589 |
| High | 5 (20%) | 24 (15.79%) |  |  |
| Normal | 20 (80%) | 128 (84.21%) |  |  |
| **Time of onset to surgery** |  |  | 238.286 (29.933–1896.887) | 2.325E-07 |
| ≥24h | 24(96%) | 14 (0.09%) |  |  |
| <24h | 1 (4%) | 138 (90.79%) |  |  |
| **Aneurysm location** |  |  | 0.927 (0.771–1.115) | 0.419 |
| PCOA | 2 (8%) | 31 (20.39%) |  |  |
| ACOA | 4 (16%) | 24 (15.79%) |  |  |
| PCA | 4 (16%) | 16 (10.53%) |  |  |
| MCA | 5 (20%) | 25 (16.45%) |  |  |
| ACA | 4 (16%) | 21 (13.81%) |  |  |
| ICA | 2 (8%) | 14 (9.21%) |  |  |
| AChA |  | 3 (1.97%) |  |  |
| BA | 2 (8%) | 7 (4.61%) |  |  |
| SCA |  | 2 (1.32%) |  |  |
| No-angiography/negatives | 2 (8%) | 9 (5.92%) |  |  |
| **Aneurysm type** |  |  | 2.38 (0.457–12.398) | 0.303 |
| No saccular aneurysm | 2 (8%) | 1 (0.66%) |  |  |
| Saccular aneurysm | 21 (84%) | 143 (94.08%) |  |  |
| No-angiography/negatives | 2 (8%) | 8 (5.26%) |  |  |
| Aneurysm size |  |  | 0.952 (0.604–1.501) | 0.834 |
| <3mm | 6 (24%) | 32 (21.05%) |  |  |
| 3–7mm | 14 (56%) | 83 (54.61%) |  |  |
| 7–10mm |  | 16 (10.53%) |  |  |
| >10mm | 3 (12%) | 10 (6.58%) |  |  |
| No-angiography/negatives | 2 (8%) | 11 (7.24%) |  |  |
| Hunt–Hess scale |  |  | 0.21 (0.097–0.458) | 0.000085 |
| Grade 1 | 16 (64%) | 60 (39.47%) |  |  |
| Grade 2 | 3 (12%) | 46 (30.26%) |  |  |
| Grade 3 | 3 (12%) | 30 (19.74%) |  |  |
| Grade 4 | 3 (12%) | 12 (7.89%) |  |  |
| Grade 5 |  | 4 (2.63%) |  |  |
| GCS scale |  |  | 0.088 (0.04–0.195) | 2.3414E-09 |
| 12–15 points | 11 | 110 |  |  |
| 11–14 points | 14 | 33 |  |  |
| 9–11 points |  | 33 |  |  |
| 3–8 points |  | 9 |  |  |
| <3 points |  |  |  |  |
| Fisher CT grade |  |  | 0.192 (0.103–0.357) | 1.7915E-07 |
| Grade 0 | 25 (100%) | 9 (5.92%) |  |  |
| Grade 1 |  | 20 (13.16%) |  |  |
| Grade 2 |  | 55 (36.18%) |  |  |
| Grade 3 |  | 41 (26.97%) |  |  |
| Grade 4 |  | 27 (17.76%) |  |  |
| MMSE scale |  |  | 0.051 (0.016–0.159) | 2.87E-07 |
| <27 points (CI) | 4 (16%) | 120 (78.43%) |  |  |
| 27–30 points (Normal) | 21 (84%) | 32 (21.57%) |  |  |
| MoCA scale |  |  | 0.039 (0.011–0.139) | 5.32E-07 |
| <26 points (CI) | 3 (12%) | 118 (77.13%) |  |  |
| ≥26 points (Normal) | 22 (88%) | 34 (22.87%) |  |  |
| EBES scale |  |  | 0.131 (0.052–0.331) | 0.000016 |
| Grade 0 | 25 (100%) | 31 (20.39%) |  |  |
| Grade 1 |  | 47 (30.92%) |  |  |
| Grade 2 |  | 41 (26.97%) |  |  |
| Grade 3 |  | 23 (15.13%) |  |  |
| Grade 4 |  | 10 (6.58%) |  |  |
| Surgical modality |  |  | 0.264 (0.092–0.756) | 0.013 |
| Endovascular coiling | 22 (88%) | 110 (72.37%) |  |  |
| Surgical clipping | 1 (4%) | 36 (23.68%) |  |  |
| Mix |  | 3 (1.97%) |  |  |
| No treatment | 2 (8%) | 3 (1.97%) |  |  |

### Univariate and multivariate regression & LASSO analysis

We performed univariate analysis of 16 indicators. Among them, smoking, time of onset to surgery, Hunt-Hess grade, GCS scale, Fisher CT grade, EBES scale, were significantly associated (p<0.05) with CI prognosis in the a-SAH group compared to the control group (Table S1).

In the training set, univariate regression identified independent risk factors associated with CI in the MoCA scale group including time of onset to surgery (OR=3.27, p=0.002), Hunt-Hess grade (OR=1.972, p=0.001), GCS scale (OR=5.601, p=0.004), Fisher CT grade (OR=2.118, p=0.000009), and EBES scale (OR=2.634, p=0.000003) (Table S1.1) In the validation set, univariate regression identified independent risk factors associated with CI in the MMSE scale group including smoking (OR=0.443, p=0.041), time of onset to surgery (OR=3.209, p=0.003), Hunt-Hess grade (OR=1.85, p=0.002), GCS scale (OR=5.111, p=0.006), Fisher CT grade (OR=2.172, p=0.000008), and EBES scale (OR=2.585, p=0.000006) (Table S 1.1). According to the ROC curves, the EBES scale had the largest AUC (AUC=0.836, p < 0.0001; Figure S1), with a clinically relevant cutoff at 1.5 points.

Multivariate logistic regression analysis confirmed that endovascular coiling showed a smaller increase in CI compared to surgical clipping in both the MoCA scale groups (B=0.239, 95%CI =0.064–0.89, p=0.033) and MMSE scale groups (B=0.198, 95%CI=0.05–0.782, p=0.021). This suggests endovascular coiling surgery had a protective effect on patients’ cognitive function. On the contrary, surgical clipping was associated with a larger increase in CI (B=4.185, 95% CI=1.123–15.594, p=0.33) indicating that patients treated with the craniotomy approach had a higher risk of CI than those that underwent endovascular coiling.

To avoid overfitting, LASSO regression screening was further applied (Figure S2, S3). We utilized ten-fold cross-validation to select the penalty term, lambda. In total, 6 variables were selected, and a Cox regression model was established based on the variables screened by Lasso regression.

### Measures of LASSO and ROC Curve Analysis

Data from the MoCA scale group (training set) was further screened using LASSO regression analysis. This yielded five factors including Hunt-Hess grade, EBES scale, GCS scale, Fisher CT Grade, and time of onset to surgery (Table 3) which were subsequently included in the logistic regression analysis below.

**Table 3.** The AUC, Cut-off, sensitivity, specificity from the ROC curve.

|  | AUC |  | 95% CI |  | Cut-off |  | Sensitivity (%) |  | Specificity (%) |  |
| --- | --- | --- | --- | --- | --- | --- | --- | --- | --- | --- |
|  | MoCA | MMSE | MoCA | MMSE | MoCA | MMSE | MoCA | MMSE | MoCA | MMSE |
| SEBE–<br>HCNNSS | 0.735 | 0.725 | 0.66–0.81 | 0.647–0.802 | 0.5 | 0.5 | 0.907 | 0.894 | 0.375 | 0.378 |
| Hunt–Hess<br>Grade | 0.651 | 0.635 | 0.566–0.736 | 0.547–0.782 | 2.5 | 2.5 | 0.318 | 0.318 | 0.083 | 0.089 |
| EBES scale | 0.744 | 0.739 | 0.669–<br>0.819 | 0.547–0.782 | 1.5 | 1.5 | 0.519 | 0.515 | 0.167 | 0.156 |
| GCS scale | 0.622 | 0.617 | 0.537–<br>0.707 | 0.53–0.704 | 1.5 | 1.5 | 0.302 | 0.295 | 0.063 | 0.067 |
| Fisher CT<br>Grade | 0.715 | 0.721 | 0.633–<br>0.796 | 0.638–0.803 | 2.5 | 2.5 | 0.473 | 0.47 | 0.146 | 0.133 |
| Time of onset to<br>surgery | 0.61 | 0.61 | 0.512–<br>0.708 | 0.509–0.710 | 0.5 | 0.5 | 0.845 | 0.841 | 0.625 | 0.622 |

The ROC values of the logistic regression are depicted in Figure 2 for the models SEBE-HCNNSS, Hunt-Hess grade, GCS score, Fisher CT grade, and EBES score. In the training set (MoCA scale group) the ROC values are 0.735(0.66–0.81), 0.651(0.566–0.736), 0.622(0.537–0.707), 0.715(0.633–0.796) respectively.

**Figure 2.**
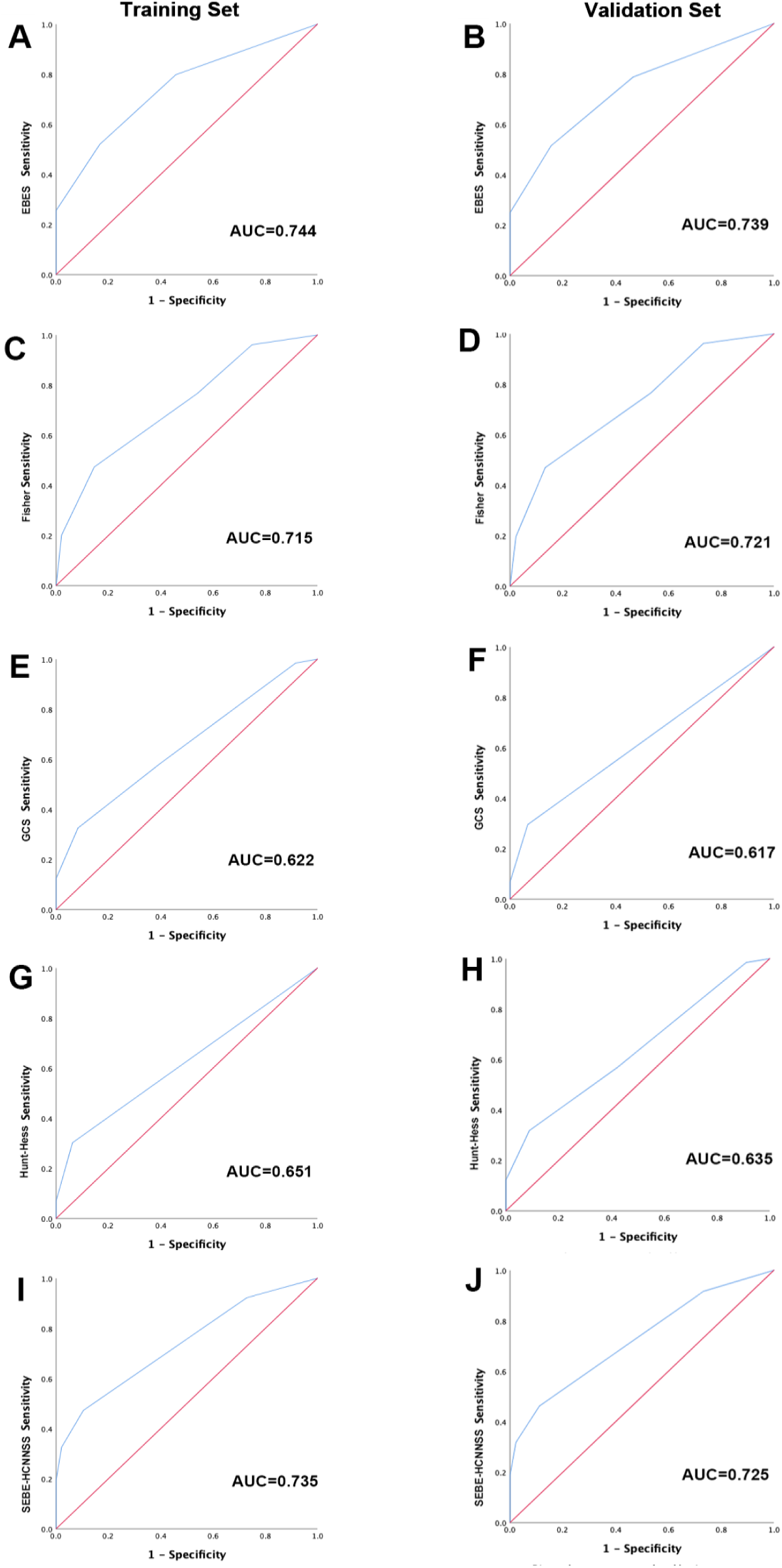
Receiver-operating characteristic (ROC) curve for prediction of unfavorable outcome and according to the clinical and radiographic grades. For all variables, SEBE-HCNNSS and the EBES scale provided essentially the same maximum area under the curve (AUC).

In the validation set (MMSE scale group) the ROC values are 0.744(0.669–0.819), 0.725(0.647–0.802), 0.635(0.547–0.7823); 0.617(0.53-0.704), 0.721(0.638–0.803), and 0.739(0.663–0.815) respectively.

The calibration test produced S:P values for the models SEBE-HCNNSS, Hunt-Hess Grade, GCS scale, Fisher CT Grade, EBES scale in the training set and validation set of 4.162, 4.403, 3.534, 3.301 and 4.548, 4.794, 3.108, 3.928, respectively. The resulting S:P values for these models in the training and validation sets indicated the robustness and validity of the models.

The corresponding p-values for the Hosmer-Lemeshow test for the five models in the training set were 0.921, 0.261, 0.416, 0.134, and 0.674. In the validation set, the corresponding p-values for the four models were 0.897, 0.144, 0.502, 0.103, and 0.658, respectively. Notably, p-values for all five models exceeded 0.05, indicating excellent model fits and validity. The DCA decision curve indicated threshold probabilities of models SEBE-HCNNSS, Hunt-Hess Grade, GCS scale, Fisher CT Grade, and EBES scale in the validation set of 44.94% to 99.13%, 50.86% to 95.73%, 68.77% to 98.29%, 40.15% to 93.73%, and 51.64% to 97.94%, respectively (Figure 3). These models demonstrated varying thresholds across a wide range of probabilities, emphasizing their utility in predicting CI (Figure 3).

**Figure 3.**
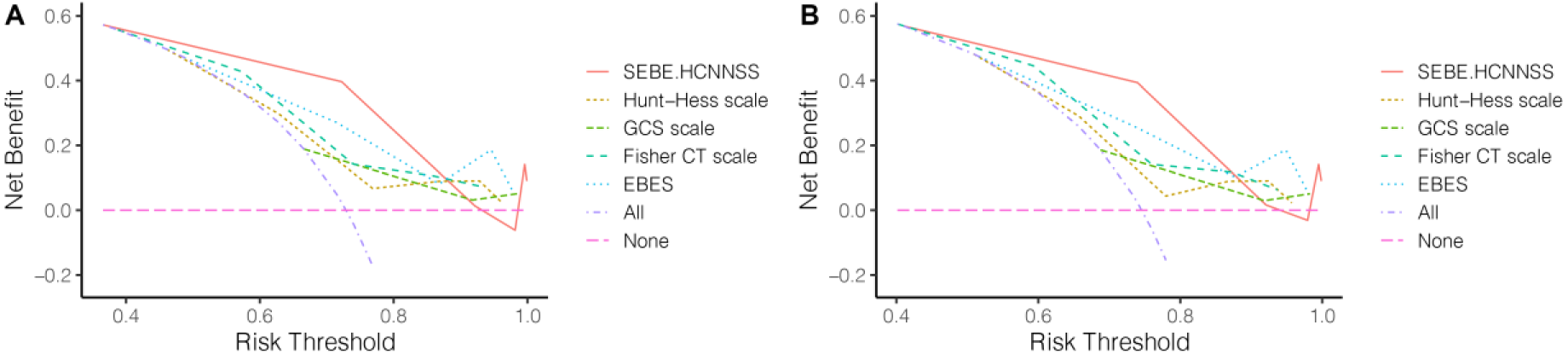
DCA curve for validating the clinical utility of the various scale. Left: DCA curve for SEBE-HCNNSS, Hunt-Hess Grade, GCS scale, Fisher CT Grade, and EBES scale in the training cohort. Right: DCA curve for SEBE-HCNNSS, Hunt-Hess Grade, GCS scale, Fisher CT Grade, and EBES scale in the validation cohort.

### Correlations between SEBE-HCNNSS and Neural Fiber Bundles, and Identification of Independent Predictors

When evaluating intracranial pressure (ICP)-related complications in SAH, such as aneurysm re-bleeding, vasospasm, and ICP elevation (Figure 4, Table S2), the SEBE-HCNNSS was dichotomously assessed after determining a clinically relevant threshold derived from the receiver-operating characteristic (ROC) curve. Variables with p<0.05 in univariate analyses were included in a stepwise multivariate logistic regression model to identify predictors of CI after a-SAH. The final model indicated notable findings: ACoA=10.492/24.295, PCoA=7.079/13.234, and 3–7mm Aneurysm =162316.229/4503.604 in the training/validation set respectively (Table S2). This suggests that changes in arterial location and aneurysm diameter was associated with an increased risk of CI, potentially attributed to compression of cerebral functional network fiber bundles. Additionally, the GCS scale was an independent predictor of CI (B=3.773, 95%CI=1.221–11.659, p=0.021)/(B= 3.51, 95% CI=1.101–11.194, p=0.034).

**Figure 4.**
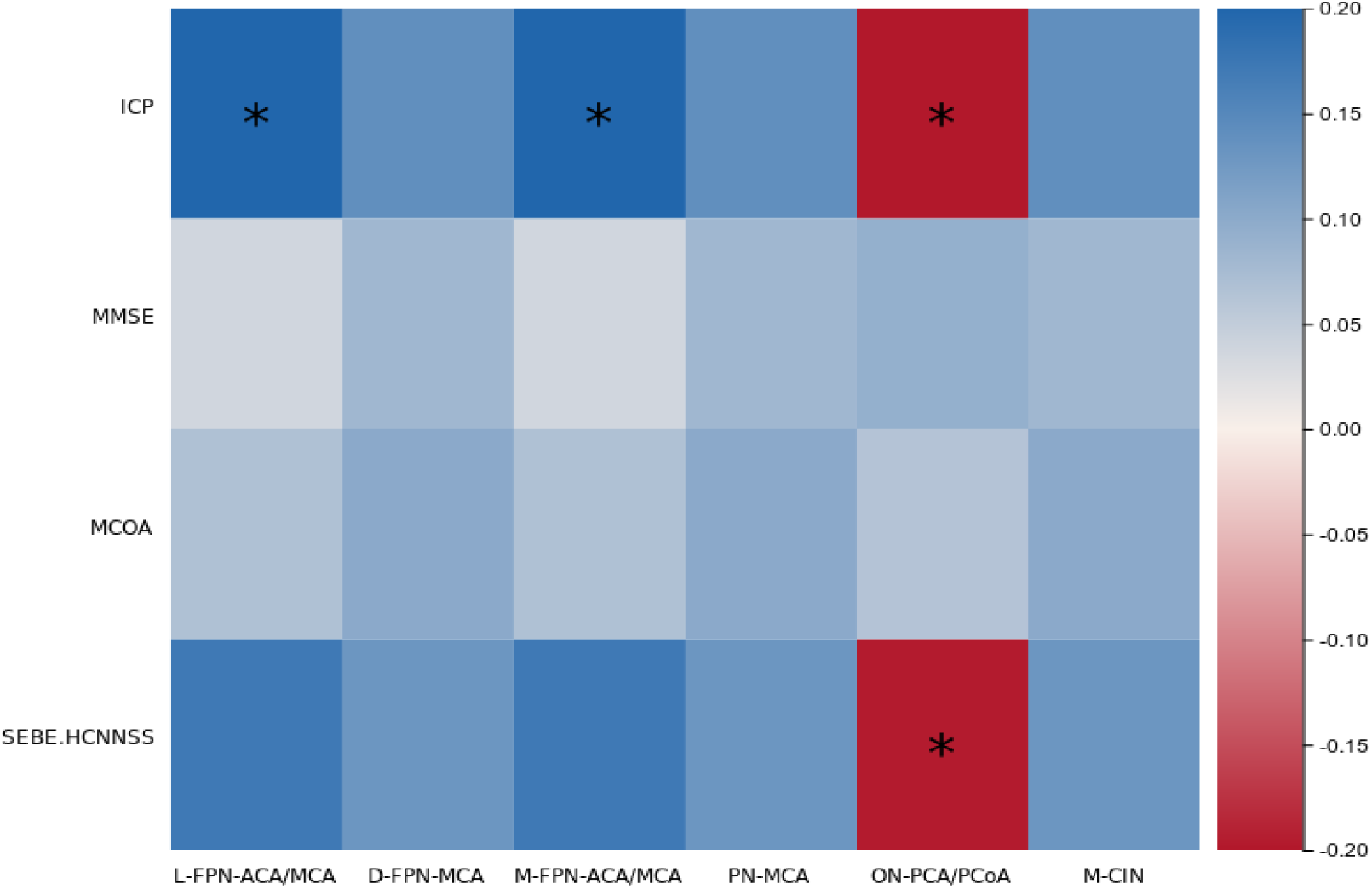
Correlation of location of arterial anatomical location and compressed nerve fibres tracts with SEBE-HCNNSS as well as MMSE and MOCA scoring models, the presence of cognitive deficits in high and low MMSE and MOCA scores correlated weaker with intracranial location than with SEBE-HCNNSS. ICP was negatively correlated with PCA r=-0.162, p=0.031; SEBE-HCNNSS was significantly correlated with aneurysm located in PCA compressing nerve fibre tracts presenting with cognitive impairment r=-0.161, p<0.05

Further analysis revealed a negative correlation between GCS scale and cognitive impairment scores in MMSE and MCOA (Person correlation=-0.185, p=0.042; Person correlation=-0.204, p=0.025). Notably, the SEBE-HCNNSS score was positively correlated with increased intracranial pressure, with higher intracranial pressure resulting in a higher rating and a correlation coefficient of 0.874 (Figure 4).

### Evaluating the SEBE-HCNNSS scale in contrast to traditional scale

The utility of SEBE-HCNNSS may be evaluated with cost-effectiveness studies^23^, supported by empirical evaluations of the impact of using the scale in clinical practice (Figure 5). Determination of net classification improvement using continuous variables in the training set established a cutoff of 1.9 (1.3523–2.448). SEBE-HCNNSS outperformed Fisher CT Grade, GCS, and Hunt-Hess Grade, while EBES scale showed superiority over SEBE-HCNNSS. Fisher CT Grade outperformed GCS and Hunt-Hess Grade. In the validation set, the cutoff was determined to be 1.7 (0.863–2.536). The EBES scale was superior to SEBE-HCNNSS, and SEBE-HCNNSS showed better performance compared to GCS scale, Hunt-Hess Grade, and Fisher CT Grade (Table 4).

**Figure 5.**
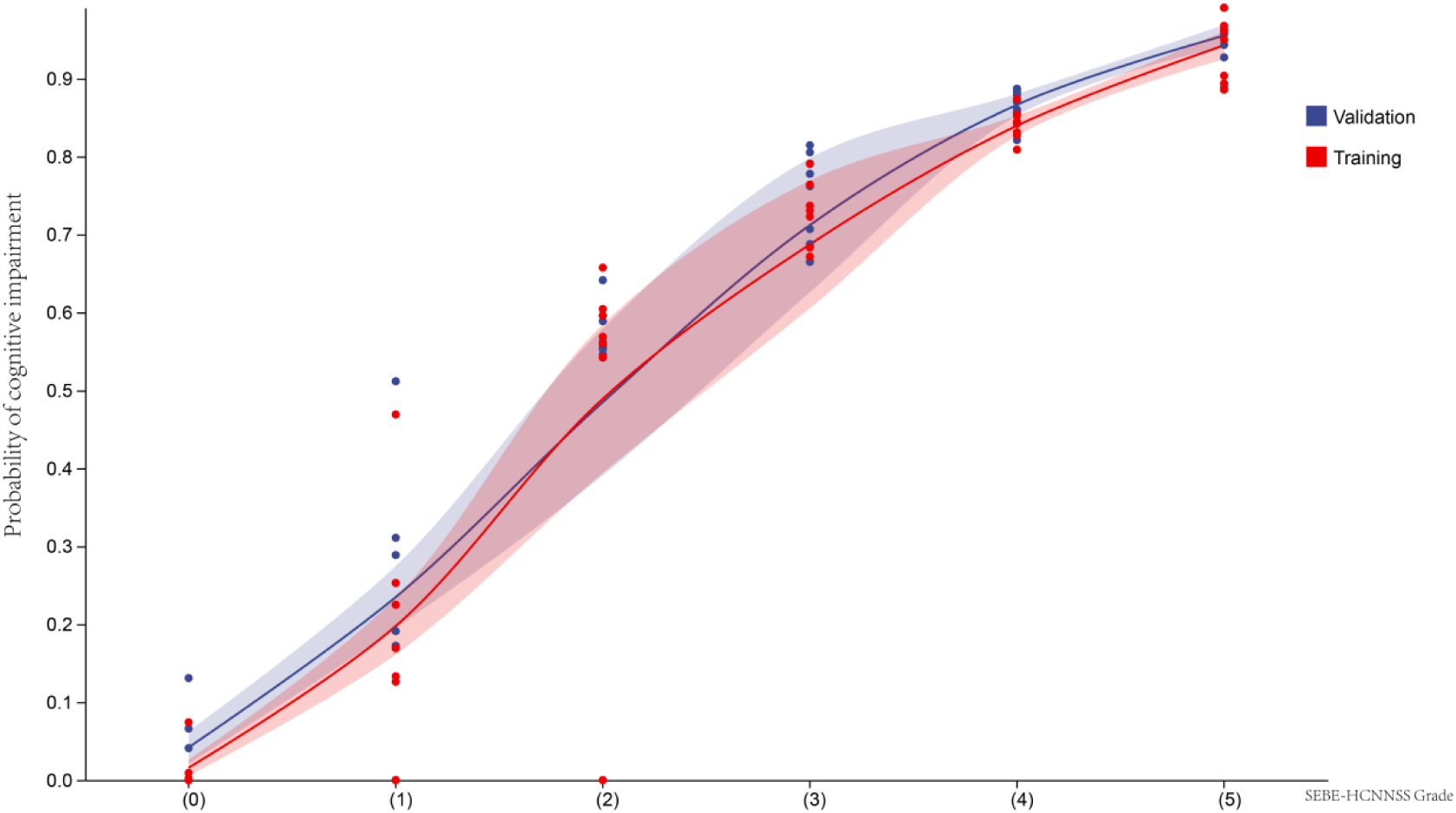
A comparison of the incidence of cognitive impairment in train <u>(MoCA scale group)</u> and validation <u>(MMSE scale group) sets</u>.

**Table 4.**
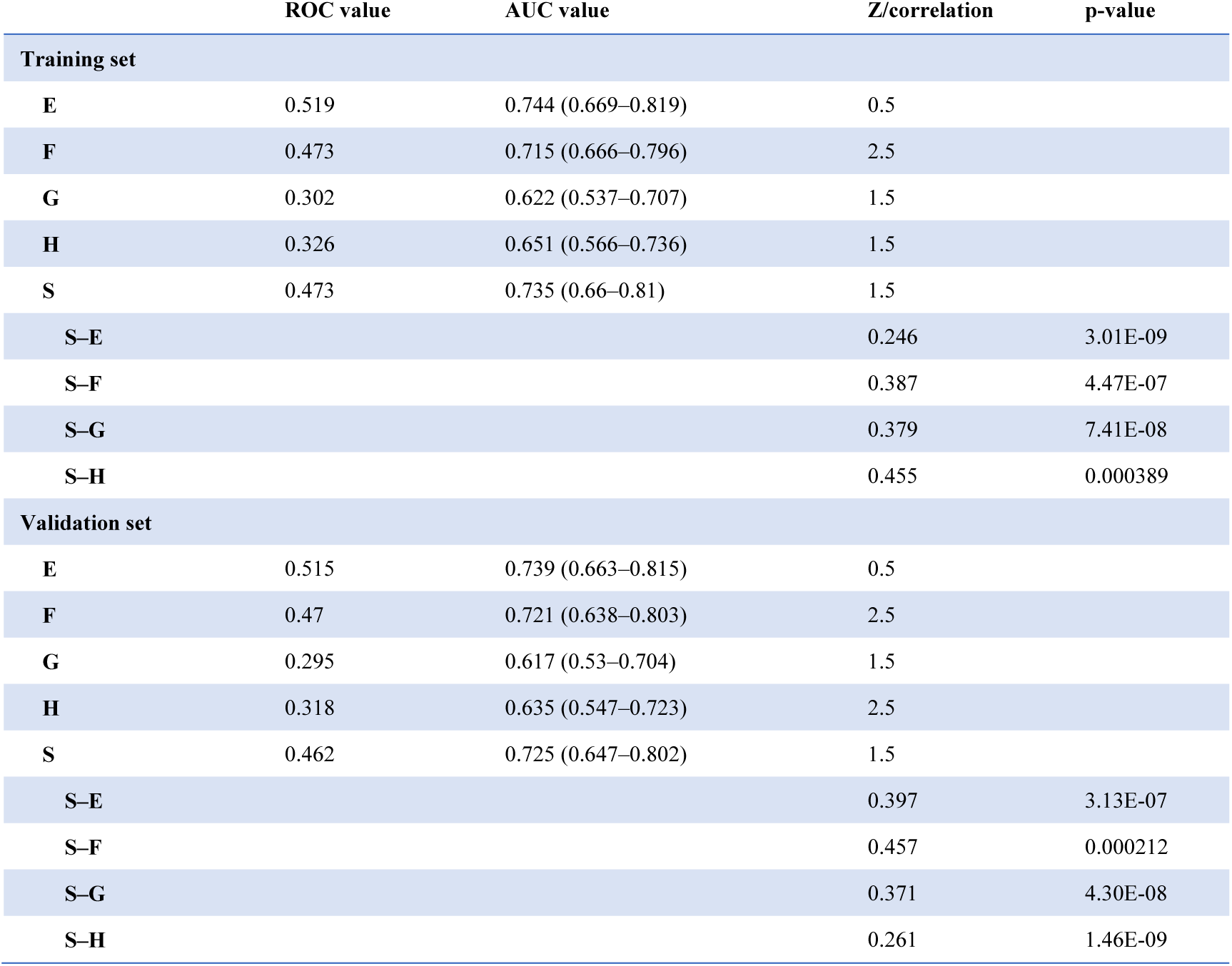
Comparison of ROC among five model.

|  | ROC value | AUC value | Z/correlation | p-value |
| --- | --- | --- | --- | --- |
| **Training set** |  |  |  |  |
| E | 0.519 | 0.744 (0.669–0.819) | 0.5 |  |
| F | 0.473 | 0.715 (0.666–0.796) | 2.5 |  |
| G | 0.302 | 0.622 (0.537–0.707) | 1.5 |  |
| H | 0.326 | 0.651 (0.566–0.736) | 1.5 |  |
| S | 0.473 | 0.735 (0.66–0.81) | 1.5 |  |
| S–E |  |  | 0.246 | 3.01E-09 |
| S–F |  |  | 0.387 | 4.47E-07 |
| S–G |  |  | 0.379 | 7.41E-08 |
| S–H |  |  | 0.455 | 0.000389 |
| **Validation set** |  |  |  |  |
| E | 0.515 | 0.739 (0.663–0.815) | 0.5 |  |
| F | 0.47 | 0.721 (0.638–0.803) | 2.5 |  |
| G | 0.295 | 0.617 (0.53–0.704) | 1.5 |  |
| H | 0.318 | 0.635 (0.547–0.723) | 2.5 |  |
| S | 0.462 | 0.725 (0.647–0.802) | 1.5 |  |
| S–E |  |  | 0.397 | 3.13E-07 |
| S–F |  |  | 0.457 | 0.000212 |
| S–G |  |  | 0.371 | 4.30E-08 |
| S–H |  |  | 0.261 | 1.46E-09 |

### Prognostic Value of SEBE-HCNNSS for CI and Unfavorable Outcomes

Among the cohort, 121 patients (68.36%) developed incident CI after SAH, and were treated by either endovascular coiling or surgical clipping or both. Notably, 43 patients (24.29%) with CI were classified as high-grade SEBE-HCNNSS, while 134 (75.71%) patients with CI were classified as low-grade SEBE-HCNNSS (Table S4). The incidence of CI increased linearly with increased severity of the SEBE-HCNNSS and edema, especially in patients exhibiting both high-grade SEBE-HCNNSS and vasospasm (p <0.05, Figure 5). The AUC of the ROC curve of SEBE-HCNNSS for the prediction of CI, edema was 0.735(0.660-0.810) and 0.725(0.647-0.802) for training and validation sets respectively, which was comparable to the AUC of the Hunt-Hess Grade, EBES, Fisher CT Grade, and GCS models. Moreover, the prognostic value of the SEBE-HCNNSS scale in predicting CI remained significant regardless of treatment modalities, including clipping (r=0.181) and coiling (r=-0.178).

### Predictors for Prognostic Value of SEBE-HCNNSS

The CT interobserver reliability of the SEBE-HCNNSS scale demonstrated a high Kappa value of 1, indicating strong agreement between observers. Among the cohort, 43 patients (24.29%) were identified with a high-grade SEBE-HCNNSS (scoring 3 to 5 points). Analysis showed that individuals with high-grade SEBE-HCNNSS were more often female (p<0.001) and exhibited higher frequency of diabetes (n=11, p=0.062), hypertension (n=22, p=0.285), smoking (n=9, p=0.912), high Hunt-Hess grade (>3points, n=17, p=0.020), high Fisher grade (>3points, n=28, p=0.000025), and high EBES scale (>3points, n=22, p<0.0001) (Table S4).

Finally Cox proportional Hazards regression was performed to identify independent predictors of CI (Table S3). In the MMSE scale group (validation set), Fisher CT Grade (OR 2.172, 95% CI 1.544-3.056, p = 0.000008), EBES scale (OR 2.585, 95% CI 1.713-3.899, p = 0.000006), GCS scale (OR 1.027, 95% CI 0.997-7.82, p = 0.051), SEBE-HCNNSS scale (OR 3.322, 95% CI 2.312-7.237, p = 0.00025) were identified as predictors of CI.

## DISCUSSION

Our study demonstrated that the SEBE-HCNNSS scale was positively associated with increased intracranial pressure following EBI after aneurysm rupture(Figure 4). Moreover, there was a direct correlation between the pathological changes induced by EBI and distinct regions of white matter fibers. Notably, the SEBE-HCNNSS was superior to traditional scales in its predictive capability, despite differences in the focus of these scales. The development of the SEBE-HCNNSS scale emerges as a more reliable predictor for EBI and CI, establishing itself as an independent prognostic indicator.

Despite the attention paid to EBI, the potential advantages of early assessment of nerve fiber tracts have often been overlooked in clinical practice^24,25^. Classification systems typically adhere to established principles within a given field. Our analysis identified predictors of high-grade SEBE-HCNNSS, including Fisher CT Grade, EBES scale, and GCS scale. In addition, our findings demonstrate varying aneurysm diameters and imaging changes in ACoA or PCoA regions are associated with an increased risk of CI. This is consistent with the pathophysiology of compression of nerve fiber bundles due to cerebral edema and hematoma lesions and in line with previous literature^26^. More specifically, the SEBE-HCNNSS grading may reflect the cascade of events post-extravasation of blood into the subarachnoid space, resulting in a sudden rise in intracranial pressure, subsequent decline in cerebral perfusion pressure, impaired autoregulation, and consequential damage to cerebral grey/white matter, microvasculature, and compression of nerve fiber bundles under increased pressure^27^. Under this model, brain anatomy may represent a physical buffer that must be depleted before the critical threshold for clinical expression of CI is reached^28^.

Furthermore, our results showed that SEBE-HCNNSS exhibited similar or enhanced predictive performance to traditional scales for unfavorable outcomes, edema, and CI (Figure 4). This is important as lesion studies suggest cognitive processes including attention, language, and memory rely on distributed processing within “multi-focal neural systems” rather than specific anatomical sites^29^. Indeed, neuroimaging highlights structural brain abnormalities as crucial risk factors for diabetic dementia, cerebral microvascular or macrovascular of diabetic complications ^30^, Alzheimer’s disease^31^, metabolic dysfunction^32^, Cerebral small vessel disease^33^ and fetal alcohol spectrum disorders^34^.

SEBE-HCNNSS is particularly valuable compared to simple cognitive exams as cognitive performance can show substantial short-term fluctuations within persons^35^. Currently, CI is increasingly recognized as a clinically significant complication in many diseases that affects the domains of memory, executive function, and language^36^. In a-SAH the increased risk of CI is especially associated with abnormal compression of brain structures with the initial severity being a strong determinant of secondary complications and functional outcome^37,38^. Thus, SEBE-HCNNSS represents a valuable prognostic indicator for a-SAH from the perspective of macro-scale functional brain networks.

In summary, our findings highlight the promising predictive value of the SEBE-HCNNSS scale following aneurysm rupture. Further comprehensive investigations and larger-scale studies are warranted to establish its broader applicability and clinical utility.

## LIMITATIONS

The study has several limitations. First, varying quality of CT scans at different medical centers, and treatment variability may have affected accuracy of collected data and definition of function outcome. Second, although the Hunt-Hess Grade, EBES, Fisher CT Grade, and GCS are commonly used outcome rating scales, some degree of observer bias is inevitable. Third, all CT scans were evaluated by a single investigator. Fourth, although interobserver agreement of the EBES scale has been found to be good, interobserver variability in grading the amount of blood may have affected accuracy of the SEBE-HCNNSS scale.

## CONCLUSION

Our findings demonstrate once again that cerebral edema and hematoma after early brain injury have no visible grooves in the initial CT slice, confirming and calculating using the SEBE-HCNNSS scale established on the basis of the SEBES grading as an important predictor of CI and poor prognosis after SAH. Elucidating the anatomical imaging mechanisms by which the pathophysiology of EBI can lead to CI, the application of imaging techniques to characterize EBI after SAH will help in the development of relevant therapeutic strategies to halt the course of brain injury after SAH and improve prognosis.

## Data Availability

none

## Acknowledgements

The authors thank the patients with SAH in the included studies and all the researchers.

## Contributors

All the authors listed above have been involved in substantial contributions to interpretation of data, conception, and design, drafting the article and revising it critically for important intellectual content and final approval in drafting the article, revising acquisition of data, and analysis of data. All authors were involved in drafting the article and reviewed and approved the final manuscript.

## Ethics approval

Institutional review board, University Medical.

## Provenance and peer review

Not commissioned; externally peer reviewed.

